# “It’s the Art of Listening”: Patient and Practitioner Perspectives on Acute Cardiac Care for Patients with a Severe Mental Illness (SMI): A Qualitative Interview Study

**DOI:** 10.64898/2026.11.12.26367223

**Authors:** Amanda Vettini, Caroline Jackson, Stewart W Mercer, Kelly Fleetwood, Sandosh Padmanabhan, Daniel Smith, Debbie Cavers

**Affiliations:** School of Population Health Sciences, Usher Institute, University of Edinburgh, Edinburgh, UK; School of Cardiovascular and Metabolic Health, University of Glasgow, Glasgow, UK; Centre for Clinical Brain Sciences, University of Edinburgh, UK; Strathclyde Institute of Pharmacy and Biomedical Sciences (SIPBS), University of Strathclyde, UK

**Keywords:** myocardial infarction, severe mental illness, acute cardiac care, quality of care, qualitative

## Abstract

**Background:** Patients with a severe mental illness experience disparities in acute cardiac care, yet reasons underlying these inequities remain poorly understood. We aimed to explore patient and healthcare professional perspectives of acute cardiac care to identify factors influencing care quality and equity.

**Methods and results:** We conducted semi-structured videocall interviews with people with a severe mental illness who had been hospitalised following a myocardial infarction, and with Cardiology and Emergency Room healthcare professionals involved in acute cardiac care across two central Scottish health boards between July 2023 and April 2024. Interviews were audio-recorded, transcribed verbatim and analysed using reflexive thematic analysis in NVivo. We conducted 21 interviews (13 healthcare professionals; 8 people with a severe mental illness). Although many patients reported empathic and supportive care, barriers were described. We identified 3 themes from the data: positive care experiences; cross-cutting care barriers, and challenges along the care pathway. Healthcare professionals sought to provide equitable care but reported challenges related to limited mental health training, diagnostic uncertainty, behavioural management and difficulties accessing psychiatric expertise, prescribed medication and associated information. Patients described experiences of stigma, communication difficulties, disruptions to psychiatric medication continuity and distress associated with overstimulating hospital environments. System-level barriers included staff shortages, limited access to liaison psychiatry, lack of clear care protocols and difficulties integrating mental and physical healthcare. Positive experiences were associated with empathic staff, feeling listened to, clear communication, and tailored efforts to create psychologically safe care environments.

**Conclusion:** Inequities in acute cardiac care for people with a severe mental illness appear to arise from the interaction of patient complexity, clinician uncertainty and organisational constraints. Improving care will require greater integration of mental and physical health services, continuity of psychiatric treatment, psychologically-informed care environments, and support for staff delivering care to this complex patient group.

**What Is Known:**

- Cardiovascular disease is a leading cause of premature mortality in people with a severe mental illness (SMI), who die up to 10-15 years earlier than the general population.
- SMI is associated with lower receipt of guideline-informed care standards for myocardial infarction (MI), which may partly explain poorer outcomes in this sub-population.

**What the Study Adds:**

- Our study provides in-depth qualitative insights on experiences of patients with a SMI and healthcare professionals (HCPs) involved in acute cardiac care, identifying barriers to equitable care.
- We identified challenges including communication, stigma, staff and medication resource deficits, patient information sharing and adequate mental health training.
- We propose recommendations for easily implementable, yet impactful, adaptations to care environments and truly listening to patients with SMI, together with improving HCPs’ mental health training, psychiatric medication access and developing tailored care protocols to improve care experiences.

## Introduction

People with a severe mental illness (SMI) have a higher incidence of cardiovascular disease (CVD), including myocardial infarction (MI), than the general population (1–3). They experience poorer MI outcomes, including a higher risk of further vascular events and higher mortality at 30-days, 1-year and 5-years (4–7). These disparities persist after adjusting for sociodemographic and clinical variables (5, 7). Despite having an increased risk of poorer outcomes, people with SMI are less likely to receive guideline-directed procedures, including coronary revascularisation, or be referred for cardiac (5, 8, 9). In a recent study where we examined multiple MI care indicators, we found that, compared to people with SMI, those with an SMI were generally less likely to receive guideline-indicated care for MI rehabilitation (10).

Qualitative insights from existing literature on hospital healthcare for people with SMI suggest multifactorial causes for care disparities at three key levels. Patient-level such as delays in help-seeking and poor self-esteem (11–13), practitioner-level such as stigma towards people with SMI (11, 14) and system-level such as poor mental-physical health services’ integration (11, 14) and overstimulating Emergency Room (ER) hospital environments with lengthy wait times (15–17). There is a lack, however, of qualitative research specifically on acute MI care experiences of patients with SMI. Our previous study with Primary Care Physicians reported challenges around prescribing cardioprotective medication to patients with SMI due to insufficient time and funding, workforce deficits, medication concordance concerns, patient complexity and difficulties navigating boundaries with mental health services (18); such challenges could also apply in the acute MI care setting.

Some progress has been made in raising awareness of the cardiovascular care issues for patients with a SMI (19), yet significant gaps in understanding remain. Whilst differences in MI care have been observed, no previous qualitative study has investigated these differences. This study aimed to address this by qualitatively exploring the perspectives of both people with a SMI who experienced hospital care for MI and healthcare professionals (HCPs) involved in delivering MI care across two geographical settings, to identify insights into relational, practical, and systemic challenges and better understand why acute MI care differences exist.

## Materials and Methods

The COnsolidated criteria for REporting Qualitative research (COREQ) were used to guide our reporting (20) and our COREQ checklist is attached (see supplementary material - Appendix 1).

### Study Design

This study forms the qualitative arm of a larger multi-method investigation, with the accompanying quantitative component using linked electronic health records to compare post-MI receipt of guideline-informed care and mortality by mental disorder and investigate whether any differences in care or mortality changed during or following the COVID-19 pandemic (4, 10). This complementary qualitative study explored experiences of HCPs across the acute cardiac care pathway and people with SMI who had experienced an MI of delivering and receiving acute MI care.

### Setting, Participants and Recruitment

The study was conducted in two urban health board regions in Scotland in the United Kingdom (UK) (Health Boards A and B). To get a range of diversity in our sampling we selected two major Scottish central belt Health Boards which both include a mix of high and low socioeconomic deprivation. People were eligible if they had a hospital admission for a suspected MI for which they were able to recall their care. We also initially defined eligibility as possessing at least one of the following SMI diagnoses: schizophrenia spectrum disorders, bipolar affective disorder, or recurrent major depression. Due to challenges in recruiting people with SMI, however, we expanded to allow inclusion of other severe mental health conditions such as severe anxiety, PTSD and dissociative personality disorder, not typically included within the umbrella of SMI. Patients were purposively sampled and recruited through the Mental Health Research Network, the Primary Care Network, and SHARE (the Scottish Health Research Register and Biobank), by displaying recruitment posters in relevant hospital outpatient departments and via professional personal networks.

HCP participants were purposively sampled to include diverse roles and experiences, including ambulance services, emergency departments, acute care cardiology units and mental health services. HCPs were eligible if they had recent experience in delivering cardiac or emergency care to patients with SMI. HCPs were recruited via clinical and research networks, newsletters circulated through professional organisations (including the Scottish Ambulance Service, Royal College of Nursing, and Royal College of Physicians), and targeted professional connections. No participants dropped out of the study once recruited. The recruitment period for both sets of participants was from May 2023 to February 2024.

### Interviews

Semi-structured interviews were carried out between July 2023 and April 2024 with no subsequent interviews occurring. The interviews were conducted by AV, a female, highly experienced, qualitative health services postdoctoral researcher with a background in public health and a PhD in sociology, employed as the Research Fellow on the study. Patient interview topic guides were developed in liaison with a patient with lived experience of SMI (see Appendix 2). Topics included patients’ health conditions, previous hospital care experiences, previous MI and their experiences along all stages of the acute cardiac care pathway. No prior relationship was established with participants prior to study commencement. Participants were informed that the researcher was a qualitative health researcher employed on the study and interested in understanding experiences of acute cardiac care.

Patient interviews were conducted primarily by NHS video call software (NearMe), with one patient opting to be interviewed face-to-face. Patients were invited to have a family member / carer present for support if they wished, with two female participants opting to have their partners present. HCP interviews were conducted remotely primarily via video call, with one HCP preferring to be interviewed via the telephone. These interviews covered: length of service; role description; comfort level in, and experiences of, caring for patients with SMI along all stages of the acute cardiac pathway; challenges to care delivery and any COVID-19 impacts (see Appendix 3).

Topic guides were iteratively refined based on developing findings and all interview data was included in the analysis. Interview questions were provided to participants in advance upon request and a small number of HCPs asked for these, however, no patients did. Interviews lasted between 35 and 80 minutes. Interviews were audio-recorded, transcribed verbatim, and de-identified. Field notes were taken by the researcher during the interview as an aide memoire to facilitate following up particular statements.

### Data Analysis

We analysed data thematically using Braun and Clarke’s six-phase reflexive inductive approach of familiarisation and quotation selection, keyword selection, coding, theme development, conceptualisation (interpreting keywords, codes and themes) and concept model development (21). Initial codes were generated inductively, and emerging patterns were refined into themes iteratively. A subset of 20% of the interview transcriptions was double coded by two experienced qualitative researchers, AV and DC, to aid discussion and conceptualisation of developing themes. Following the double coding process, further refinements were made to the coding framework, analysis and themes. Data were managed using NVivo software. Analysis included comparison across patient and HCP interviews and involved regular reflexive memos and discussions by researchers to consider assumptions about the participants and the data and ensure a transparent and robust analysis. Analysis of patients’ data focused on the patient’s account, while carers contributed contextual information during interviews. Themes were developed to reflect both convergent and divergent perspectives within and across patient and HCP interviews, looking for patterning across the data and, later, noting where this converged with existing literature. No formal coding tree was produced because analysis used reflexive thematic analysis; Figure 1 presents the final thematic structure.

Data collection ceased when sufficient depth and diversity of experiences had been captured to support development of robust themes across both participant groups; it is inappropriate to refer to data saturation within Braun & Clarke’s methodology. Transcripts were not typically returned to participants for correction although we contacted one HCP to clarify an element of their transcript.

### Ethical Considerations

Ethical approval for the study was granted by the NHS Research Ethics Committee (REC - 22/WS/0141). All participants were given a participant information sheet and patients had at least a week to consider the information before providing consent. At least 12 months must have elapsed since hospitalisation for their MI prior to interview and Mental Health (MH) clinicians only referred those patients considered sufficiently psychiatrically stable. GPs were informed of patients’ participation in our study. Participant anonymity has been preserved by using numeric identifiers. All participants, including HCPs, were given a shopping voucher as thanks for their participation.

## Results

We conducted 21 interviews: 13 with HCPs and eight with people with SMI who experienced MI. Participant characteristics are described below.

### Characteristics of people with an SMI

People with SMI (six male, two female) had a range of severe mental health conditions: schizophrenia, severe depression, severe anxiety, post-traumatic stress disorder (PTSD) and dissociative personality disorder. We were unable to recruit any patients with bipolar disorder. Using Harrison et al’s definition, half of our patients had comorbidity (22), disclosing one or more other chronic conditions during their interview when prompted. Patients’ characteristics are presented in Table 1. Both female patients opted to be accompanied by their partners during the interview, who were also interviewed in their capacity as a carer. All patients were in Health Board A and there was variability in the extent of contact with health services regarding number and length of hospital stays.

### HCPs’ Characteristics

Thirteen HCPs were interviewed: nine from Health Board A and four from Health Board B. These were paramedics, nurses and doctors working in a variety of roles across the MI care pathway, in different settings and with a range of experience levels (see Table 2).

### Findings

Our analysis produced three core themes: 1. Positive care experiences, 2. Cross-cutting care barriers and 3. Care challenges at acute cardiac care pathway stages (see Figure 1 for core themes and sub-themes). Table 3 summarises key problems in care delivery and some proposed solutions.

Findings below are structured according to these core themes and sub-themes. Each quote includes the participant group (patient, denoted by ‘P’, or HCP), their participant number and further key information of SMI for patients where S = schizophrenia and D = depression for patients and job role for HCPs where N = nurse, MHN = mental health nurse, D = doctor and P = paramedic for HCPs. People with SMI will be hereafter referred to as ‘patients’. Participants frequently referred to mental health rather than SMI which is notable in many quotations.

### Theme 1: Positive care experiences

Good practice and positive care experiences were described by patients, praising staff for their dedication and exemplary treatment of them.

*The staff are lovely…they work hard*. (P6, D)

*Everybody was dead nice. I just ended up being really embarrassed about everything.* (P2, S)

### Empathic care

Increased empathy for patients manifested via tender actions was remarked upon by both patients and HCPs as critical in improving their care experiences. A patient spoke of a particularly impactful and emotive empathic care experience.

*A…nurse… said, are you alright? I said, I don’t know if I’m going to live tomorrow. She actually gave me a cuddle**…** I just sat there crying with her.* (P4, D)

A sense of clinicians going above-and-beyond typical care standards emerged from both participant groups. An ER nurse reported making additional effort for with patients with SMI.

*I almost give more to…regular attenders, which often fall into the severe mental illness category, and that’s because I always think, what is so wrong in their life that they’re constantly coming in? (*HCP 11, N)

A patient expressed feeling positively overwhelmed by their heart bypass Consultant’s care in accompanying them to a procedure in different hospital being undertaken by another surgeon.

*They [the Consultant] said, don’t worry I’m coming with you in case they need me. And that was absolutely amazing of him. There’s not many consultants who’ll actually change their whole schedule to look after one patient*. (P4, D)

### Clear information provision / reassuring staff demeanour

Clear pre-procedural explanations and staff demeanour exuding confidence and levity helped to relax and reassure patients.

*They explained what they were going to do [in the angiogram], what they were looking for…They were quite funny, so they made you feel relaxed.* (Patient 6, D)

*I felt confident because they were confident.* (P7, D)

HCPs identified a heightened need for information among this patient group. To meet this need, some HCPs took additional care by keeping patients well-informed seeking to avoid unnecessary distress or by providing pre-procedural visits.

*When people have mental health…. Keeping them well informed, it’s just trying to prevent a trigger*. (HCP 8, N)

*If someone has a mental health disorder I would always see them myself… beforehand… to try and give them some reassurance.* (HCP9, D)

Others such as this ER Consultant took additional time to listen attentively.

*You have to listen to them. You have to take time.”* (HCP12, D)

### Creating safe environments

To create safe, nurturing care environments, some HCPs enhanced efforts at patient familiarisation and environmental tailoring, providing reassuring routines gleaned from information from families and patients to develop a comfortable atmosphere.

*Get a routine that is comfortable with the patient…makes all the difference… If their family member can tell you that they like their blind halfway up rather than all the way up**…**it helps them relax more and become more comfortable to receive care.* (HCP11, N)

A doctor ruminated on whether clinicians could go further in seeking to provide the kind of environment that would help patients feel more relaxed.

*I just wonder whether there’s anything that we could do better as clinicians to try and make them feel more comfy and at ease.* (HCP10, D)

Considering optimal placement beside other patients within ER so as to not unnecessarily distress this patient group was a strategy that some HCPs employed.

*Patient placement in the department is key…You don’t put them in…a cubicle opposite some others who are drunk, shouting and screaming.* (HCP13, N)

### Theme 2: Cross-cutting care barriers

Several cross-cutting barriers to positive care experiences were found including: difficulties managing complex /multimorbid patients; overall challenges well-integrating physical and mental healthcare; challenging patient behaviour; resources deficits of people or things (for example, staffing shortfalls compounded by intensive workloads); the distressing nature of the hospital environment for some, and medication stock and access problems disrupting continuity. Barriers around communication and attitudes also featured: stigma; uncertainty in caring for patients with SMI; diagnostic overshadowing; problems accessing MH teams, and deficits in information sharing / patient records access. Lack of training and protocols for MH also posed problems.

### Comorbidity / complexity management challenges

Acute cardiac care appeared to operate sufficiently well for patients in our study with only one psychiatric condition, especially if well-managed. Problems, however, sometimes emerged caring for those with comorbidity on multiple medications, with those patients reporting the poorest quality cardiac care.

*If I’m on your ward and you’re a leg doctor and I have a leg problem, you’re a happy person because you know how to fix that… But if I come in as a complex case, forget it.* (P4, D)

A patient with comorbidity reported being incorrectly administered naloxone, an opioid overdose reversal treatment.

*I remember them giving me some form of injection, and then the pain, I was screaming… the consequence of that [naloxone injection] was I’d taken another one [heart attack.]… I am on a lot of medication, and back then, I was on a lot more. And they probably thought I was a junkie.* (P6, D)

*They presumed….because she’s on extensive medication…a lot of it is opiate based… So, they jumped to the conclusion that she’d taken an overdose…They injected her with naloxone.* (P6 Carer, D)

Multiple negative experiences in hospital had a layering effect with some comorbid patients recounting feeling alienated from, and not trusting, healthcare.

*I’m not looking for honours, I’m just looking to be treated like a member of society that has at least some value. And now I feel as if I’m a nobody, I’m a number.* (P4, D)

### Challenges integrating physical and mental healthcare

Patients and HCPs identified difficulties integrating mental and physical healthcare within hospital MI settings. One patient perceived that delays in accessing psychiatric medication were not afforded the same urgency as would be expected for a physical health medication such as insulin, reflecting concerns about the parity of esteem afforded to mental and physical healthcare needs.

*“I just think it is strange. If I had diabetes, they wouldn’t stop my insulin.”* (P2, S)

HCPs similarly described uncertainties and divisions in attempting to meet both the physical and mental healthcare needs of patients with SMI within a system primarily designed around acute physical illness.

*I think within healthcare maybe it’s seen as you’re either a physical health nurse or you’re a mental health nurse, and some people just don’t want to know. (*HCP7, N)

At times practitioners felt discomfort from feeling unable to deliver the quality of holistic care they aspired to provide.

*Sometimes you walk away from the end of the shift thinking that you’ve not been able to provide that person with the best care possible. (*HCP7, N)

### Patient behaviour difficulties

Many staff and some patients spoke of concerns about challenging, distressing and sometimes aggressive behaviour of patients with SMI they had experienced due to SMI and psychosis.

*I’ve been strangled once by a patient… He was having an acute psychotic episode.* (HCP9, D)

*I started getting quite verbally abusive to the nurses because I thought I had been kidnapped*. (P2, S)

## Resource deficits

### Staffing shortfalls and intensive workloads

Staffing constraints were a recurrent concern, particularly in relation to supporting patients with SMI who typically required more time, supervision, or relational care. HCPs wished to show empathy and take additional time but lacked capacity, noted by HCPs and some patients.

*We are all hit by…the challenges of staffing…quite often are short staffed so that can obviously impact care.* (HCP8, N)

*Most of them do tell you [procedural explanations]…But sometimes they’re otherwise pre-occupied. They might be thinking about other things, and they just want to get their job done.* (P5, D)

A psychiatric nurse reported that the need for several of their staff members to accompany their patients to other hospitals, often for extended periods, placed pressure on staff levels within their psychiatric hospital.

*That does have a huge impact on us down at the [psychiatric] hospital.* (HCP1, MHN)

### Distressing hospital environments

Patients typically arrive at ER to be assessed before being appropriately triaged elsewhere in the hospital. The lack of psychologically-informed environment of ER and cardiac wards with bright lighting, high noise levels and crowded spaces was described as particularly distressing for patients with psychosis, PTSD, or anxiety. Such environments can exacerbate agitation, being a catalyst for challenging behaviour and impact HCP’s ability to build trust and give the care needed.

*A patient…that was quite manic at the time was…running around the department and was very agitated… because of the noise and the light, getting more and more worked up*. (HCP10, D)

*One patient…who had a diagnosis of schizophrenia who was a STEMI (ST Elevated Myocardial Infarction) was in our coronary care unit….It was quite challenging to build up a trusting relationship with this person because they were quite paranoid and having delusions during their presentation.* (HCP7, N)

*This patient that I’m thinking of in particular, I suppose you [patients] feel like you’re in a bit of a goldfish bowl as well, with people looking at you.* (HCP7, N)

Long wait times and shared spaces exacerbated feelings of paranoia or fear, with patients and HCPs recounting instances of absconding or early self-discharge.

*A patient with severe depression…severe anxiety and PTSD who found being in a shared bay incredibly stressful to the point… they discharged themselves before they were medically fit to do so.* (HCP7, N)

*The absconding is running away. That’s the PTSD kicking in* (P4, D)

Sometimes side rooms are used as a solution to the overstimulating hospital environment, but these are neither routinely available nor embraced by all patients with SMI, as demonstrated by the following contrasting responses.

*I got bumped into a room on my own and I hated that…It was quite isolating, and it gives you a lot of time to think.* (P7, D)

*I got a room to myself for a while… I liked it very much.* (P2, S)

### Medication – Stock, access and impacts

Some HCPs reported the crucial importance of psychiatric medication continuance in avoiding triggering mental health deterioration.

*When we’re seeing these people, we need to make sure that they’ve got their medications and they’re getting them regularly and on time. Because if they even miss like one or two doses, that could send them into some sort of acute mental health deterioration.* (HCP 10, D)

Nonetheless, HCPs acknowledged delays in medication access and highlighted system-level issues such as limited ward stock and unclear prescribing responsibility.

*Something very niche…one or two days and they’ve not had their medication…we’re trying to get it sourced.* (HCP3, D)

Problems accessing medication impacted negatively on HCPs, some of whom spent lengthy periods attempting to source or verify medication.

*That takes time to phone a pharmacy, to corroborate what they’re on. It can take an hour of my time to do that.* (HCP12, D)

Some described discomfort at being unable to access psychiatry records for dosage and timing information, and reliance on pharmacy and liaison psychiatry, who were not always available.

Such delays could exacerbate psychiatric symptoms and increase the risk of deterioration or behavioural disturbance, as well as patients reporting feeling abandoned.

*I think my antipsychotic was starting to dry out of my system. I did mention I had schizophrenia at that point…Nobody seemed to take that seriously, surprisingly…I ended up having a psychotic episode in the ward* (P2, S)

### Mental health training – deficit and advice-seeking

HCPs in all three role types raised deficiencies in their formal training for managing mental illness within acute cardiac settings. Many described feeling underprepared to engage with people with SMI exhibiting challenging behaviour.

*As nurses, we do not have a vast knowledge regarding mental health and how to deal with people when their behaviour’s challenging… It’s not something that we see on a daily basis. (*HCP 8, N)

This often led to reliance on ‘common sense’, previous exposure, or informal peer learning. Some staff expressed anxiety and discomfort around psychiatric symptoms feeling inexperienced.

*I think a lot of it is nurses might be quite afraid because they don’t know how to deal with [patients with a SMI]…you know, and they might feel they lack experience to know what to say as well.* (HCP 2, N)

One paramedic conveyed severe deficiencies in their MH training, whilst the other had received some, but found it challenging to apply:

*I have had zero training in mental health in my whole career, absolutely none.* (HCP4, P)

*We have had some training in mental health…but it is over such a wide range of conditions, everyone presents differently.* (HCP6, P)

Most doctors also indicated limited MH training and solved this by seeking advice from psychiatric staff.

*I often will not feel confident to do that and you go straight to the mental health team for advice…* (HCP3, D)

Applying training at the pace required in a busy ER department was challenging, even when MH training was perceived as adequate.

*We did have quite a bit…But actually on the job, it’s completely different…especially in A&E (ER) when you see these patients really quickly.* (HCP10, D)

### Communication and information provision / sharing barriers

Both HCPs and some patients identified care barriers relating to communication and information sharing, including stigma towards patients, inconsistency in care plan provision, and a lack of clear protocols for caring for people with SMI.

#### Stigma and uncertainty

Widespread stigmatisation towards mental illness amongst physical healthcare staff was frequently commented upon by both patients and acknowledged by HCPs. Some patients felt treated as an inconvenience, dismissed as unworthy of equivalent attention as patients without psychiatric conditions.

*I was starting to get voices and stuff and I asked for help a couple of times. The nurse actually said to me ‘Doctors are busy seeing sick people’…I don’t think they take it seriously enough, to be honest*…*: I was really angry.* (P2, S)

A lack of effective MH training played into HCPs stigma towards mental illness, creating fear and uncertainty in handling this patient group.

*People being wary of people with mental health…There’s that…fear factor…that affects the way in which people approach someone.* (HCP7, N)

A psychiatric nurse concurred regarding cardiology practitioners’ fear:

*They [cardiology nurses] do seek a lot of support from us, with our patient load, because I think there is a level of fear there.* (HCP1, MHN)

#### Lack of clear mental health protocols and care plans, insensitive information sharing, and challenges accessing MH teams’ support

HCPs described a lack of formal protocols for managing psychiatric emergencies within cardiac settings. While there were clear guidelines for responding to medical deterioration, the approach to escalating behavioural or mental health crises was often ad hoc. In some cases, this led to the use of physical restraint or sedatives.

Y*ou have to… restrain patient in A&E, take bloods in order to…to actually find out if something happened there.* (HCP 13, N)

Several participants expressed a desire for clearer guidance, improved training, and stronger links to mental health crisis teams to manage these situations more safely and ethically.

*If they become very agitated… there’s no clear protocol in A&E (ER) …for anything else, you have protocols.* (HCP13, N)

Access to care plans replete with valuable contextual information was critical in providing good patient care, although not consistently available for all.

*If they’ve [patients with SMI] seen a few people before about their mental health, they usually have…a care plan… Not everybody had that but that was actually quite useful.* (HCP10, D)

Systemic issues meant that potentially useful information was not always routinely shared with HCPs. A paramedic highlighted that they were not always informed of mental health status prior to emergency call attendance; some potentially relevant information such as prior aggression was typically linked to addresses not individuals.

*I wouldn’t [be MH status informed] unless… the patient had been verbally abusive in the past or… an incident at that address… obviously that’s not just anyone with a mental health problem…And it’s not attached to the person, it’s attached to the address.”* (HCP 4, P)

Some patients were disconcerted by HCPs’ information delivery and/or conduct. One patient overheard their attending paramedic saying the patient might die and the other paramedic appeared visibly distressed.

*The two ambulance guys, I guess it was their first time seeing a heart attack victim because I thought he was going to burst out crying…And then the other guy I could hear him outside the ambulance on the radio and he said ‘look we’re gonna need a police escort or he’s not gonna make it.* (P7, D)

Another patient received a disquieting angiogram risk explanation.

*The consultant… told me about the angiogram thing and he… frightened me a little bit… He said that the risks…’There’s one in five thousand chance of you dying through this test… you could be one of the unlucky.’* (P5, D)

### Theme 3: Barriers along the acute cardiac care pathway

Systemic, patient-related and practitioner-related barriers exist at each stage of the acute cardiac care pathway from the initial call for help, arrival at ER, gaining a patient history, investigative tests, heart procedures, discharge to post-admission aftercare.

### Delayed help-seeking /symptom minimisation

Some patients described hesitating to seek medical help during the onset of cardiac symptoms, interpreting their symptoms as minor or unrelated to cardiac health instead attributing them to anxiety, indigestion, or panic. The emotional barrier of not wishing to be a burden to anyone was a recurring theme.

*And me thinking it was just indigestion that I had, I thought I don’t want to bother anyone. …I just jumped on the bus*. (P5, D)

*I didn’t feel bad enough to call an ambulance*. (P2, S)

### Gathering a patient history and investigative tests

Most clinicians spoke of patients’ difficulties in opening up to give a history, some patients could be ‘withdrawn’ due to their mental health and they also communicate ‘differently’ to other patients.

*You get patients that are quite withdrawn that don’t necessarily want to communicate…much.* (HCP8, N)

*They might… describe it differently.* (HCP2, N)

HCPs described challenges they faced in interpreting patients’ communication regarding symptoms and events leading up to the suspected MI to be able to form an adequate picture to enable diagnosis. Information was frequently piecemeal and not provided simultaneously.

*It was quite difficult to get a grip on a chain of events*. (HCP2, N)

Diagnostic overshadowing, whereby a patient’s physical problem is incorrectly attributed to their mental health condition, was reported to potentially affect diagnosis:

*General view of mental illness…an assumption that anything the patient is presenting with will be related to their mental health.* (HCP12, D)

### Procedural challenges and care differences

Regarding decisions about whether to perform diagnostic tests such as an angiogram, some clinicians discussed whether an agitated patient could remain sufficiently still for the procedure.

*It’s a procedure that you need to lie flat for, for at least an hour. If this patient is showing a degree of agitation…would need to weigh up the risks versus the benefits… for angiograms.* (HCP8, N)

HCPs also highlighted challenges of contrasting situations whereby some people with SMI refuse investigations such as blood tests due to suspicion / mistrust, yet others proactively request this.

*People…that had schizophrenia were quite suspicious that we were taking bloods for other reasons… In a few situations, we couldn’t get bloods, it was too dangerous.* (HCP10, D)

*It can almost go the other way where you don’t think they needs bloods done but they’re demanding bloods. So that can be difficult because if you set a precedent for always taking bloods with people who are…regularly coming back.* (HCP11, N)

Many HCPs expressed concerns about patients’ behaviour often leading to procedural modifications. Concerns about post-operative medication adherence resulted in departing from standard practice, such as not inserting stents during angioplasties.

*Committing someone to a stent that requires regular anticoagulant drugs…could be catastrophic if he had a lapse in his mental health condition and decided not to take any medication. Bypass surgery would be safer in that setting.* (HCP9, D)

HCPs reported that patients with SMI required more reassurance and information that other patients, in one case necessitating continuous explanation during a cardiology procedure.

*[Patient who had a] small heart attack, and a significant mental health disorder…was extremely anxious…Every procedure he wanted to talk through as we did it in real time…A very difficult scenario because I need to concentrate on doing the procedure.* (HCP9, D)

### Discharge challenges

Both HCPs and patients commented on difficulties in discharging patients with more complex SMIs and needs, resulting in them having to remain in the cardiology hospital ward.

*A guy who had a severe mental illness and there wasn’t a clear pathway for him and he ended up being an inpatient for months and months.* (HCP7, N)

Length of hospital stay seemed to negatively impact care too; patients who had lengthy stays reported feeling like a burden to hospital staff. One patient reported feeling resented by staff.

*Although it’s against NHS policy, I was being called a bed-blocker…you could hear some people talking about you.* (P4, D)

#### Concerning discharge

A patient reported a concerning discharge of being permitted to travel home unaccompanied on public transport a few hours post angioplasty and stent, despite post-operative dizziness.

*I: Did someone come and get you and take you home?*

*R: No… I don’t have a car and she [patient’s sister] doesn’t drive, so I just got on a bus.* (P5, D)

### Other findings – Non-complaining

A minor theme around some patients with SMI not wishing to complain despite dissatisfaction was noted.

*But I think maybe he [the doctor] should’ve chosen his words a bit more carefully [when explaining the risks of an angiogram]. But I didn’t say anything because I don’t like to get anyone into trouble on an official thing.* (P5, D)

*No, I was never going to complain about something like that, you know the state of the NHS, at lease I wasnae sitting in a corridor. (Patient 7)*

## Discussion

The central finding of this study is that inequities experienced by patients with SMI during acute cardiac care appear to arise from layered interactions between patient complexity, limited mental health expertise, systemic constraints and healthcare environments not well-configured for the needs of people with a SMI. Structural and systemic issues meant that organisational arrangements and service configurations inadvertently disadvantaged people with SMI. Examples identified in our study included lack of clear protocols, inconsistent access to psychiatric expertise, difficulties obtaining prescribed psychiatric medication and wider challenges integrating mental and physical healthcare. Difficulties obtaining prescribed psychiatric medication emerged as a particularly tangible example of the challenges created by separation of mental and physical healthcare systems.

### Comparison with existing literature

Empathy has been shown to benefit patient care both for those with SMI and without; some of our patients remarked upon the significance of empathy in positive care experiences (23–25). Notwithstanding this, our findings add to previous evidence showing that even when individual clinicians seek to provide high quality, empathic hospital care, the systemic structure constrains their capacity to do so (15, 17, 26). Our qualitative and quantitative studies’ findings mutually complement and support one another. Our quantitative study found that patients with schizophrenia, bipolar disorder or depression generally had higher mortality risk following STEMI or NSTEMI, even after adjusting for confounders, excepting for patients with bipolar disorder who showed no evidence of a difference in 30-day mortality following NSTEMI (4). We also found that reduced receipt of guideline-informed care standards amongst those with a mental disorder may partially explain lower one-year post-MI mortality (10). While our qualitative data cannot establish causality, they identify several plausible mechanisms through which disparities may arise, including diagnostic uncertainty, difficulties obtaining psychiatric information and medication, behavioural management concerns, and fragmented communication and information sharing across the healthcare service. Together, our qualitative and quantitative findings provide convergent evidence that inequalities in acute cardiac care are unlikely to be explained solely by patient factors.

Some medical clinicians hold perceptions about patients with an SMI of increased care difficulty, reduced medication and treatment adherence, and pessimism about prognosis (14, 27). Some also report limited training in mental health and difficulties obtaining procedural consent all of which we identified in our study. Such uncertainty may contribute to reliance on assumptions about patients’ behaviour, capacity, or treatment adherence, which in turn can influence clinical decision-making. Diagnostic overshadowing has traditionally been examined within emergency and general medical settings (26, 28); our findings indicate that it may also influence decision-making along the MI hospital care pathway, where assumptions about patient behaviour, symptom reporting or capacity may shape assessment and treatment trajectories. We found that interviewed clinicians reported diagnostic and procedural challenges with this patient group, sometimes resulting in opting to treat them differently. This identifies plausible mechanisms through which disparities in acute cardiac care may arise and potentially contribute to observed differences in outcomes.

Although trauma-informed approaches have received increasing attention within mental health services, they remain less visible within acute cardiac care. Trauma-informed models emphasise the importance of safety, trust, collaboration and attention to the physical care environment, recognising that healthcare encounters and overstimulating clinical settings may inadvertently exacerbate distress (15, 16, 29). Our findings suggest that trauma-informed principles may offer a useful framework for improving experiences of care among patients with SMI admitted with acute MI. It is worth recognising, however, that this is a complex and challenging patient group to care for; our previous work identified family physicians’ challenges in cardioprotective prescribing and care for patients with SMI due to their complexity (18). Previous literature has also identified that patients with chronic mental disorders report challenges accessing primary medical care (30).

Some patients appeared reluctant to challenge care decisions or formally complain about negative experiences. Although we were unable to determine the reasons for this, previous research has demonstrated associations between mental illness-related stigma, reduced self-efficacy and diminished confidence in healthcare interactions (12, 13, 31).

Perhaps most strikingly, patients often remembered how they were treated rather than what treatment they received. Positive experiences were often attributed not to specialised interventions but to relational aspects of care such as feeling listened to, respected and reassured. This suggests that improvements in patient experience may not always require substantial resource investment but can arise through comparatively small interpersonal behaviours that foster trust and psychological safety.

### Implications for practice

Our findings suggest how greater alignment of mental and physical healthcare within acute cardiac care could be achieved. Where mental health training is bolstered this can help improve care (32). Our findings underline the need for targeted mental health training for all frontline staff working in cardiac and emergency settings, which would likely alleviate the general uncertainty in managing patient’s behavioural distress in cardiac settings (16, 19). Improved access to liaison psychiatry, the creation of shared protocols for managing psychiatric crises in cardiac settings and removing any barriers to accessing mental health records and medication information through ensuring records’ integration and training for all staff on how to access them, could help address many of the system-level barriers described.

Additionally, there is a need to re-evaluate hospital through a psychologically-informed lens. Quiet spaces, patient placement, clear communication, and continuity of care can significantly reduce distress and improve outcomes (15, 16, 29). Our proposed strategies echo calls from other studies for more comprehensive care models that truly acknowledge the interplay between mental and physical health (19, 30). Our key finding of the importance of empathic care and feeling heard aligns with evidence on the crucial importance of empathy, positive attitudes and how even a solitary ‘good’ HCP can substantially transform patients’ experiences (15). This reinforces our point that seemingly small or individual-level efforts can make big differences.

### Strengths and Limitations

This study’s strengths include diversity of professional roles interviewed along the acute cardiac care pathway, the richness of data across both patient and staff perspectives and being a dual-site study. This study shares insights from an especially hard-to-reach patient group overcoming significant recruitment challenges to do so. In providing qualitative data, this study complements the results from our quantitative study (4, 10) by delivering potential explanations for its findings on care differences and increased mortality.

This was a small-scale, exploratory study. While we took steps to include participants from two different health boards to explore contextual variations, patient and HCP perspectives may not be applicable to other contexts, particularly internationally. Challenges with recruitment resulted in a smaller number of participants than originally aimed for We were only able to recruit patients from Health Board A whose healthcare experiences could potentially differ from other Scottish health boards. We did not collect socioeconomic data and were therefore unable to explore how deprivation and social disadvantage may have influenced experiences of care. Our findings may underestimate the scale of the problem. Although we recruited patients with a range of psychiatric diagnoses, we were unable to recruit participants with bipolar disorder. Furthermore, our sample appeared healthier than many people with SMI (2, 3), with lower levels of comorbidity, suggesting that the challenges identified may underestimate those experienced by patients with greater clinical complexity. Moreover, increased risk of post-MI mortality at 30-days and 1-year among this patient group (5–7) means that the sickest patients would likely have died before being able to be included in our research.

### Recommendations and implications for future work

Table 3 presents suggested strategies for addressing the barriers identified in this study and highlights opportunities for both individual-level and system-level improvement. Further larger-scale qualitative research is warranted to explore the findings from our exploratory study with a larger sample size, Future research should seek to gain the perspective of patients with bipolar disorder, who were missing from this study, and to explore in greater depth the experiences of those with complex comorbidity, which was identified as a particular care challenge. Problems both sourcing and obtaining existing prescribed psychiatric medication were reported by some of our patients and HCPs; further quantitative research to document the prevalence of this issue would be pertinent.

Future studies should also explore the perspectives of general practitioners/family physicians, community mental health teams and liaison psychiatry services. There is also scope to evaluate interventions aimed at improving medication continuity, staff training, and integrated care planning for this population. Implementation studies should investigate how liaison psychiatry and trauma-informed practices can be scaled and adapted across cardiac settings. Co-designed solutions that include patients, carers, and multidisciplinary staff are essential to ensure feasibility and equity. Ultimately, this study demonstrates the importance of listening and trying to understand the broader context of the patient’s life and mental condition as well as their physical symptoms.

Further work to explore the role of side rooms is warranted. Our evidence suggests they can be helpful for some patients, but a source of distress and isolation for others. Side rooms are also typically both in limited supply and are prioritised for infection control or restraint purposes. By attending not only to the physical symptoms of heart disease, but to the broader psychosocial context in which care occurs, healthcare systems can better meet the needs of people living with SMI. A ‘one size fits all’ approach truly does not work here and careful, individualised tailoring is required.

## Conclusions

This qualitative study highlights some good practice, but also significant and interrelated challenges faced by both patients with SMI and HCPs in the context of acute cardiac care. Addressing these challenges requires a multi-level response that includes integrating mental health and cardiology services, ensuring wards are appropriately staffed, improving staff training in mental health care, and improving systems to ensure continuity of medications and supportive care environments. Meaningful improvement will depend on policies that promote collaboration across disciplines, and that include the voices of patients and clinicians in co-designing solutions.

In conclusion, while HCPs strive to deliver equitable acute cardiac care, patients with SMI continue to encounter barriers arising from the complex interaction of clinical, organisational and environmental factors. Improving care will require not only greater integration of mental and physical health services but also increased attention to the relational aspects of care that patients valued most. As one Emergency Room nurse observed, caring effectively for people with SMI is often “*the art of listening*”, an observation that captures both the complexity of the challenge and the simplicity of one of its most salient solutions.

## Author contributions

CJ, KF, DC, DJS, SP and SWM conceived the study and were awarded funding. AV conducted the majority of the qualitative analysis supported by DC. All authors contributed to the interpretation of the results. AV wrote the first draft of the article with DC providing initial comments. All authors commented on the revised first draft and approved the final version.

## Data Availability

The qualitative interview transcriptions are deposited in Zenodo: https://doi.org/10.5281/zenodo.23586496

## Acknowledgements

The authors gratefully acknowledge the patients and healthcare professionals who participated in the semi-structured interviews, giving so generously of their time and generating such rich and valuable data. The work was carried out with the support of the Mental Health Research Network, the Primary Care Network, and SHARE (Scottish Health Research Register) who helped with participant recruitment. Additionally, the Mental Health Research Network assisted recruitment by displaying recruitment posters in relevant hospital outpatient departments, professional contacts and word of mouth. We would also like to thank the two health boards for advertising our study within their networks.

## Nonstandard Abbreviations and Acronyms

ER: Emergency Room
CVD: Cardiovascular Disease
GP: General Practitioner (Primary Health Care Physician)
HCP: Healthcare Professional
MI: Myocardial Infarction
NSTEMI: Non-ST Elevated Myocardial Infarction
SMI: Severe Mental Illness
STEMI: ST Elevated Myocardial Infarction

## References

1. Goldfarb M, De Hert M, Detraux J, Di Palo K, Munir H, Music S, et al. Severe Mental Illness and Cardiovascular Disease: JACC State-of-the-Art Review. J Am Coll Cardiol. 2022;80(9):918–33.

2. Hert M, Correll CU, Bobes J, Cetkovich-Bakmas M, Cohen DAN, Asai I, et al. Physical illness in patients with severe mental disorders. I. Prevalence, impact of medications and disparities in health care. World Psychiatry. 2011;10(1):52– 77.

3. Reilly S, Olier I, Planner C, Doran T, Reeves D, Ashcroft DM, et al. Inequalities in physical comorbidity: a longitudinal comparative cohort study of people with severe mental illness in the UK. BMJ Open. 2015;5(12):e009010.

4. Fleetwood K, Nolan J, Berry C, Cavers D, Mercer SW, Padmanabhan S, et al. Mental disorders, mortality following myocardial infarction, and the impact of the COVID-19 pandemic in England: a cohort study. Eur Heart J-Qual Car. 2026.

5. Fleetwood K, Wild SH, Smith DJ, Mercer SW, Licence K, Sudlow CLM, et al. Severe mental illness and mortality and coronary revascularisation following a myocardial infarction: a retrospective cohort study. BMC Med. 2021;19(1):67–13.

6. Chung Chang W, Kwun Nam Chan J, Sau Man Wong C, Chi Fai Or P, Siu Han Hai J. S83. Mortality, revascularization and cardioprotective pharmacotherapy after acute coronary syndrome in patients with psychotic disorders: A population-based cohort study. Schizophrenia bulletin. 2020;46(Supplement_1):S65–S6.

7. Bodén R, Molin E, Jernberg T, Kieler H, Lindahl B, Sundström J. Higher mortality after myocardial infarction in patients with severe mental illness: a nationwide cohort study. J Intern Med. 2015;277(6):727–36.

8. Chan JKN, Chu RST, Hung C, Law JWY, Wong CSM, Chang WC. Mortality, Revascularization, and Cardioprotective Pharmacotherapy After Acute Coronary Syndrome in Patients With Severe Mental Illness: A Systematic Review and Meta-analysis. Schizophrenia Bull. 2022;48(5):981–98.

9. Shao M, Zhuo C, Gao X, Chen C, Xu Y, Tian H, et al. Reduced rate of revascularization in schizophrenic patients with acute myocardial infarction: A systematic review and meta-analysis. Prof Neuro-Psychoph. 2020;99:109870.

10. Fleetwood K, Nolan, J., Berry, C., Cavers, D., Mercer, S.W., Padmanabhan, S., Smith, D.J., Stewart, R., Vettini, A., Jackson, C.A. Mental disorders, receipt of acute cardiac care following myocardial infarction and the impact of the COVID-19 pandemic: a cohort study. BMJ Open 2026.

11. Costa M, Meunier-Beillard N, Guillermet E, Cros L, Demassiet V, Hude W, et al. Results of a Qualitative Study Aimed at Building a Programme to Reduce Cardiovascular Risk in People with Severe Mental Illness. Int J Env Res Pub He. 2022;19(11):6847.

12. Brohan E, Gauci D, Sartorius N, Thornicroft G. Self-stigma, empowerment and perceived discrimination among people with bipolar disorder or depression in 13 European countries: The GAMIAN–Europe study. J Affect Disorders. 2011;129(1):56–63.

13. Olcun Z, Altun OS. The Correlation Between Schizophrenic Patients’ Level of Internalized Stigma and Their Level of Hope. Arch Psychiat Nurs. 2017;31(4):332–7.

14. Stone EM, Chen LN, Daumit GL, Linden S, McGinty EE. General Medical Clinicians’ Attitudes Toward People with Serious Mental Illness: A Scoping Review. J Behav Health Serv Res. 2019;46(4):656–79.

15. Navas C, Wells L, Bartels SA, Walker M. Patient and Provider Perspectives on Emergency Department Care Experiences among People with Mental Health Concerns. Healthcare-Basel. 2022;10(7):1297.

16. Austin EE, Cheek C, Richardson L, Testa L, Dominello A, Long JC, et al. Improving emergency department care for adults presenting with mental illness: a systematic review of strategies and their impact on outcomes, experience, and performance. Front Psychiatry. 2024;15:1368129.

17. Clarke DE, Dusome D, Hughes L. Emergency department from the mental health client’s perspective. Int J Ment Halth Nu. 2007;16(2):126–31.

18. Vettini A, Brennan GK, Mercer SW, Jackson CA. Severe mental illness and cardioprotective medication prescribing: a qualitative study in general practice. BJGP Open. 2024;8(2):1–12.

19. Borkowski P, Borkowska N. Understanding Mental Health Challenges in Cardiovascular Care. Cureus J Med Science. 2024;16(2):e54402.

20. Tong A, Sainsbury P, Craig J. Consolidated criteria for reporting qualitative research (COREQ): a 32-item checklist for interviews and focus groups. Int J Qual Health C. 2007;19(6):349–57.

21. Braun V. Thematic analysis: a practical guide. Clarke V, editor. London: London: SAGE Publications; 2022.

22. Harrison C, Fortin M, van den Akker M, Mair F, Calderon-Larranaga A, Boland F, et al. Comorbidity versus multimorbidity: Why it matters. J Multimorb Comorb. 2021;11:2633556521993993.

23. Winter R, Leanage N, Roberts N, Norman RI, Howick J. Experiences of empathy training in healthcare: A systematic review of qualitative studies. Patient Educ Couns. 2022;105(10):3017–37.

24. Hamed WE, Babegi AS, Elamin HAM, Kamel NA, Escolano-Castillo AM, Dailah HG, et al. Stigmatizing attitudes and predictors of empathy toward mentally ill patients among psychiatric and mental health nurses. BMC Nurs. 2025;24(1):454.

25. Ruffalo ML, Kottapalli M, Anbukkarasu P. Empathy in the Care of Individuals With Schizophrenia: A Vital Element of Treatment. American Journal of Psychotherapy. 2024;77(1):30–4.

26. Shefer G, Henderson C, Howard LM, Murray J, Thornicroft G. Diagnostic Overshadowing and Other Challenges Involved in the Diagnostic Process of Patients with Mental Illness Who Present in Emergency Departments with Physical Symptoms – A Qualitative Study. PLOS ONE. 2014;9(11):e111682.

27. Noblett JE, Lawrence R, Smith JG. The attitudes of general hospital doctors toward patients with comorbid mental illness. Int J Psychiat Med. 2015;50(4):370–82.

28. Liberati E, Kelly S, Price A, Richards N, Gibson J, Olsson A, et al. Diagnostic inequalities relating to physical healthcare among people with mental health conditions: a systematic review. Eclinicalmedicine. 2025;80:103026–.

29. Greenwald A, Kelly A, Mathew T, Thomas L. Trauma-informed care in the emergency department: concepts and recommendations for integrating practices into emergency medicine. Med Educ Online. 2023;28(1):2178366.

30. Levinson Miller C, Druss BG, Dombrowski EA, Rosenheck RA. Barriers to primary medical care among patients at a community mental health center. Psychiatr Serv. 2003;54(8):1158–60.

31. Hofer A, Mizuno Y, Wartelsteiner F, Wolfgang Fleischhacker W, Frajo-Apor B, Kemmler G, et al. Quality of life in schizophrenia and bipolar disorder: The impact of symptomatic remission and resilience. Eur Psychiat. 2017;46:42–7.

32. Hassan S, Heinkel S, Burton A, Blackburn R, McCloud T, Ross J, et al. A qualitative study exploring the barriers and facilitators of implementing a cardiovascular disease risk reducing intervention for people with severe mental illness into primary care contexts across England: the ‘PRIMROSE’ trial. BMC Health Serv Res. 2020;20(1):753–15.

